# Genotypic Characterization of Urinary Tract Infections causing bacteria isolates among Adults at Kiambu Level 5 Hospital, Kenya: Selected Extended Spectrum β-Lactamase genes and Biofilm Formation

**DOI:** 10.1101/2022.10.23.22281223

**Authors:** Fredrick Kimunya Wanja, Eric Omori Omwenga, Caroline Wangare Ngugi, John Ndemi Maina, John Ndemi Kiiru

**Affiliations:** Department of Medical Microbiology, Jomo Kenyatta University of Agriculture and Technology, P.O. Box 6200-00200 Nairobi, Kenya; Department of Medical Microbiology & Parasitology, Kisii University, P.O. Box 408-40202, Kisii, Kenya; Centre of Microbiology Research, Kenya medical Research Institute, P.O. Box 43640-00100 Nairobi, Kenya

**Keywords:** Antimicrobial resistance (AMR), Resistance genes, Biofilm formation, Antibiofilm inhibitory effects, Extended Spectrum β- Lactamase (ESβLs)

## Abstract

The menace of antimicrobial resistance to public health is constantly arising globally. Many pathogenic bacteria use mechanisms such as mutations and biofilm formation, which significantly reduces efficacy of antimicrobial agents. In this cross-sectional study, we aimed at determining the prevalence of selected extended spectrum β-lactamase (ESβLs) genes and analyse the possible biofilm formation abilities of the isolated bacteria causing urinary tract infection among adult patients seeking medicare at Kiambu Level 5 hospital, Kenya. The double-disk synergy test was used for phenotypic identification of ESβLs producing isolates, while microtiter plate assays with some modifications were used to test biofilm formation analysis. A total of 10 isolates were bioassayed for ESβL genes presence out of 57 bacteria isolates obtained from urine samples. From this study, the *bla*_*TEM*_ genes were found to be the most prevalent ESβLs genes (100%), followed by *bla*_*OXA*_ and *bla*_*SHV*_ genes at 40% and 30% respectively. In addition, the co-carriage of *bla*_*TEM*_ and *bla*_*SHV*_ was revealed at 50% lower than that of *bla_TEM_ + bla*_OXA_ genes at 66.7% among the study *E. coli* isolates. Biofilm formation finding disclosed that most of the isolates form biofilms 36 (63.2%), with Gram-negatives being the most biofilm formers 25 (69.4%) compared to the Gram-positive 11 (30.6%). *E. coli* 15(41.7 %), *Klebsiella sp*. 7(19.4%) and *S. aureus* 7(19.4%) were the most common biofilm formers. Further analysis showed no significant difference in biofilm formation among all tested isolates with a p-value of more than 0.05. However, overall Gram-positive isolates had a significant P-value of 0.056. Although biofilm formation’s impact on urinary tract infections is not fully recognized, the carriage of ESβLs resistance genes and the biofilm formation ability negatively impact effectiveness of UTI treatment. Therefore, we advocate for surveillance studies to map ESβLs distribution and biofilm formation genes among UTI etiological agents to halt UTI treatment failure.

## 1.0 Introduction

Bacterial UTIs continue to be a major public health concern that affects both the genders and age groups globally (1). These infections can easily be treated with existing antimicrobials, but treatment options are becoming fewer with the ever-increasing, (re)-emergence and spread of antimicrobial resistance (AMR) (2). The situation has been made worse with the emergence of multi drug and extra drug resistant strains with projections of reaching soon an antibiotic free world in the horizon (1). This has been attributed to the lack of or poor regulation implementation and widespread use/misuse of antibiotics globally; hence the resistance patterns of bacteria causing UTIs are changing drastically. This has been worsened by the emergence and spread of AMR, which has become more rampant in developing countries, Kenya included, with minimal strategies being employed to curb this menace (2). The change in AMR patterns has been attributed to various strategies being employed by bacteria, like the hydrolyzing activities of extended-spectrum β-lactamase (ESβL) and biofilm formation abilities of the UTIs causing pathogen which ends up shielding the bacteria from potent antimicrobial agents amongst many (3). For instance, the carriage of ESβLs genes by bacteria causing UTIs has been cited for mediating resistance to extended spectrum Cephalosporins (3). This is despite the fact that most β-lactam antibiotics have been reported to be potent against both Gram-positive and Gram-negative bacteria. The recent noted resistance cases against this class of antibiotics are actually of immense clinical significance (4). With this knowledge and many more, there is a pressing need to build up amicable antimicrobial stewardship to better our understanding of the changing antibiotic resistance trends.

Various studies done at the global level have documented enormous and ever-increasing distribution levels of ESβL producing bacteria, a trend that should be of great concern globally (5). For example, a study done in Iran indicated the mass prevalence of the presence of β-lactamase genes (96.3%) amongst the study isolates (6). Almost similar results were documented in a recent study conducted in the Philippines, where the prevalence and distribution of ESβLs-encoding genes was 66.6% (5). This could be attributed to the fact that ESβLs are mostly located on plasmids and transposons, which easily facilitates the transmission dynamics of these resistance traits amongst pathogens within and between species presenting an infection control risk, particularly within hospitals (7).

However, regionally, and particularly in Kenya, the knowledge of carriage of EβSLs among urinary tract infection etiological agents is still a normal challenge beyond most clinicians reach (4). In addition, the limited knowledge and the rising AMR among bacterial agents causing UTIs due to the carriage of EβSLs can be associated with the poor epidemiological surveys, phenotypic and genotypic characterization of multi-drug resistance micro-organisms in developing countries. As a result, UTIs caused by pathogens possessing EβSLs genes have proved difficult to treat due to limited antibiotic treatment options that are not readily available in these poor resource-limited settings (7).

Biofilms are exopolysaccharide polymers formed on a mass of bacterial cells that then houses them as they keep on multiplying and producing more virulence traits (8). Even though pathogenic and non-pathogenic bacteria can form biofilms, some of the most common bacteria causing UTIs have been identified to be good biofilm formers (8). Among the most common UTI causing bacterial isolates includes; *E. coli, S. aureus* and *P. aeruginosa*, even though *E. coli* has been singled out as the most notorious biofilm former (8). According to a prior study, bacteria that exist in biofilms exhibit extraordinary resistance to conventional biocides, antimicrobial treatments and host immune response compared to their planktonic (non-adherent) counterparts (9). Once micro-organisms form biofilms, they alter their phenotypic characteristics, reducing their susceptibility to antibiotics (10). This influences microbial responses to stimuli in medical and non-medical settings, especially the responses towards antimicrobial agents and the host immune system. The observed phenotypic changes are associated with the microbe genetic diversity since the expression of genes within the said community leads to biofilm formation (11). To succeed in UTI treatment and halt recurrency, more research on biofilm formation and the impact association to resistance of UTI causative agents to commonly used antibiotics will help tailor prudent treatment solutions.

This ability to form biofilms by most UTI etiological agents, the contribution of the biofilms to uropathogens resistance to antibiotics needs to be well understood. Furthermore, our understanding of the interplay between biofilm and AMR among bacteria causing urinary tract infections worldwide and how to best treat these infections is limited (10). Again, the distribution of biofilm formation genes among bacteria causing UTIs is under-investigated. However, microbes that live closely in biofilms are said to embrace cellular signaling communication mechanisms to ensure all collaboration for optimum benefits. Future studies on the association between biofilm formation and AMR should be seen as a greater way of understanding biofilm processes, leading to novel, effective antibiofilm inhibitory strategies and improving patient management (11). The clinical effect of these biofilms and how they afford pathogens a favorable environment to thrive should be seen a scientific gap that needs to be elucidated. These biofilms also give pathogens more time to escape the cidal/ static activities of antimicrobials (12)(13), meaning more genetic and molecular advances will also better our understanding of these bacterial communities, boosting treatment.

To assist halt biofilm establishment and dissolution of the already existing ones, long-term solutions with significant antibiofilm inhibitory effects are paramount (14). More so, because the interplay between antibiotic resistance and biofilm formation remains unclear and has a significant impact on the cost of medical treatment and prevention of infectious diseases (15)(16). The use of drugs with antibiofilm inhibitory effects will therefore aid in unmasking the biofilm exopolysaccharide matrix allowing efficient penetration of antibiotics and host immune responses into biofilms, which will result in minimum resistance and hampering of antibiotic therapy (17). This current study finding therefore sheds light on the prevalence of EβSLs and possible abilities to biofilm formation among bacterial isolates causing UTI among adult patients seeking medicare at Kiambu Level 5 hospital, Kenya. The revealed finding also equips health caregivers with crucial information that will diagnose and probable potential treatment solutions for urinary tract infections.

## Materials and Methods

### 2.1 Study design, site and ethical approval

This cross-sectional study was done in Kiambu level 5 Hospital, Kiambu County – Kenya (1°10′S36°50′E). The study’s ethical approval was sought from the Kenyatta National Hospital-University of Nairobi Ethical Research Committee (Reference no: KNH-ERC/A/470) and National Commission for Science, Technology, and Innovation (NACOSTI) (Reference no: 619853).

#### 2.1.1 Ethics Statement

The principal investigator was at the laboratory reception all the time and explained the goal of the study to each adult patient into details to ensure smooth recruitment exercise. After the detailed explanation, consent of the participant was sought, which required him/her to fill in and sign the consent form (Supplementary data S1).

### 2.2 Sample size

This present study involved isolates from our previous study by Wanja *et al*. (18)where 57 significant bacterial isolates and 31 non-significant growths below the ≥100,000 CFU/ml (10^5^) threshold were obtained. Therefore, the 57 UTI-positive isolates were used as our sample size to determine the possibility of biofilm formation. On the other hand, isolates resistant to 3rd generation cephalosporins (Ceftriaxone, Cefotaxime and/or Ceftazidime) and ampicillin were screened for the EβSLs genes carriage (18).

### 2.3. Molecular detection of EβSLs

#### i. Phenotypic detection of EβSLs

A double-disk synergy test to detect the likelihood of isolates carriage of EβSLs was performed using disks of 3^rd^ generation Cephalosporins and Cephalosporin-inhibitor (Clavulanic acid) antimicrobial disk as previously described (19) (20). The test disks of 3^rd^ generation Cephalosporins and Ceftazidime/Clavulanic acid were used. On inoculated Mueller-Hinton agar, the disks were kept 30mm apart, centre to centre. A clear extension of the edge of the inhibition zone of Cephalosporin towards the Augmentin disk was observed was interpreted as a positive for EβSL production as done before (18). All tests were independent of each other.

#### ii. DNA extraction

The DNA template for the PCR test was obtained using the boiling method as previously described (21). Briefly, a loopful of pure bacterial inoculum was immersed into 1mL of PCR grade water (Invitrogen, DNase/RNase free) in a 2mL Eppendorf tube. Lysis of the bacterial cell membrane was done by boiling at 95°C for 15 minutes in a thermal block (Twin incubator, DG 210). The separation of DNA from other cell extracts was done by centrifuging (BioSan LMC-3000 Centrifuge; Serial no; 90804012, CHINA) at 1400 rpm for 5 minutes. The supernatant containing extracted DNA was transferred to another Eppendorf tube and stored at -20°C (21).

#### iii. Genotypic Identification of EβSLs Genes

Molecular screening was done by PCR method to determine the carriage of selected EβSLs genes (*bla*CTX-M, *bla*SHV, *bla*TEM and *bla*_OXA_) as described by previous studies (22) (23) (24). The final volume in each PCR tube was 25µl which included 10 µl of Qiagen master mix, 1 µl butane, 2 µl DNA, 10 µl PCR water, and 2 µl forwards and reverse primer (specific to each target gene). Amplification was done using a thermal cycler (GeneAmp® PCR system 9700) under the following conditions; initial denaturation at 95 °C for 2 min, annealing at 50 - 60 °C (depending on the primer) for 1 min, extension at 65 °C for 8 min and a single final extension step at 65 °C for 8 min for 30 cycles. Amplified PCR products were separated in 1.5% gel and banding patterns were visualized under a UV gel imager.

### iv. Genetic analysis of E. coli Isolates

Resistance to either of the tested expanded spectrum cephalosporin was used as a basis for selection of six *E. coli* isolates for repetitive tandem repeats analysis. This analysis was done using the GTG 5-PCR method using published strategies and primers indicated in **Table 2.1**. PCR amplification of the target DNA sequence was done at an initial step of denaturing for about 1 minutes at 95°c, then the second fold of annealing at 40°C temperature and finally the third fold of the final extension at 72°c. Amplified products were separated by running in 1% agarose gel for I hour. Visualization of banding patterns was done using a Gelmax® UV imager. Banding patterns were analyzed using bionumerics Gelcompar®2 software versions 6.6, with the cluster analysis done using the dice method based on UPGMA arithmetic mean. As previously described, a correlation of ≥80% among bacterial species was regarded as strong evidence of genetic relatedness among isolates (25).

**Table 2.1:**
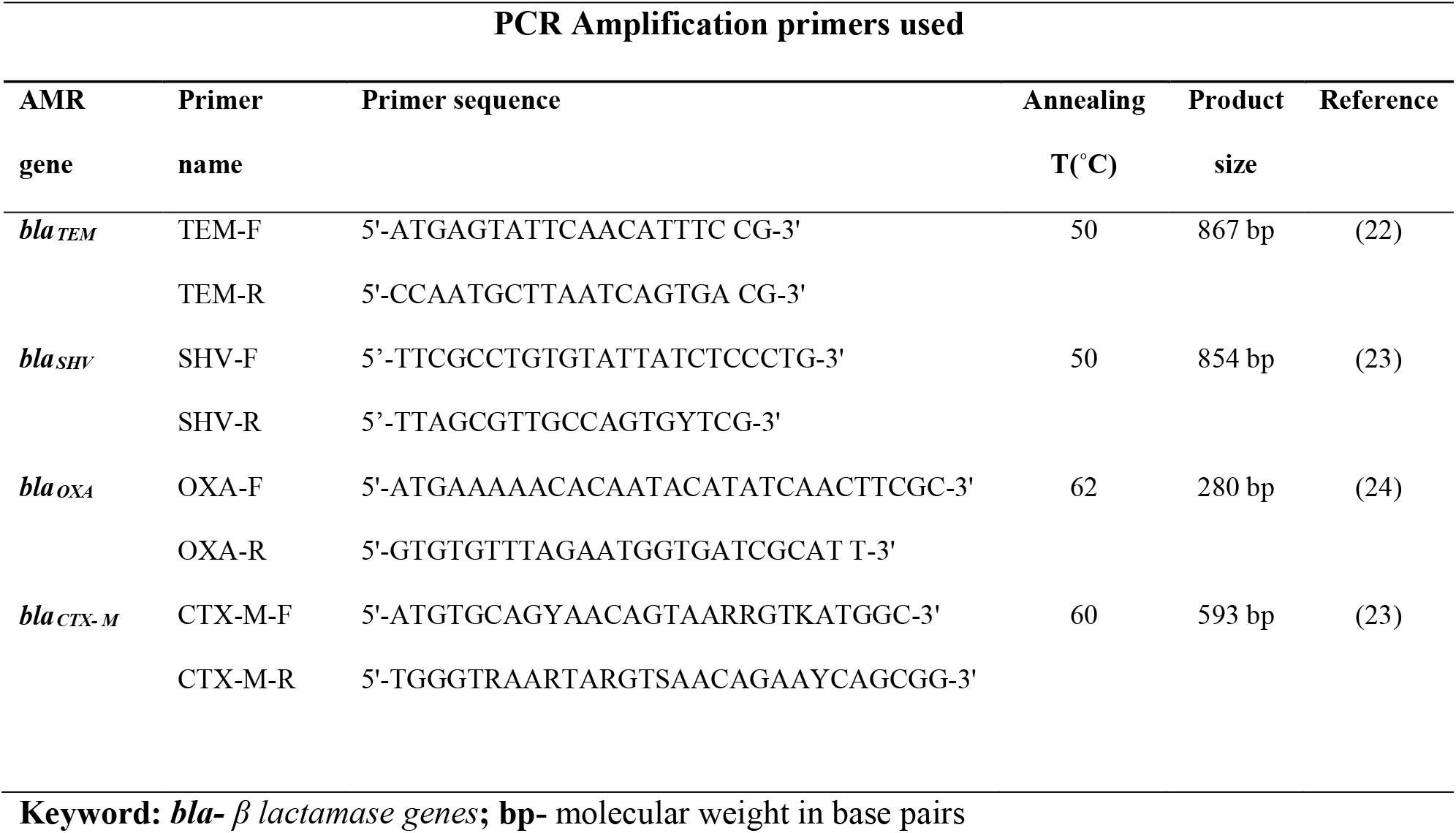
PCR Amplification primers used

### 2.4. Biofilm formation Assay

The study test isolates’ ability to form biofilm was determined using 96 well microtiter plate assays with some modifications as described before (26). The isolates stored at -80°C in vials were first thawed, 10 µl picked and inoculated into 190µl Tryptone Soya Broth (TSB-OXOID, UK) and incubated in a microtiter plate for 48 hours at 37°C without shaking. By use of a parafilm, the flat-bottomed 96 well microtiter plates were sealed for purposes of preventing medium evaporation. After the incubation, the microtiter plates were rinsed gently 2-3 times with double-distilled water 200µl to remove loosely attached cells/planktonic cells. The microtiter plate was then air-dried for 1 hour before adding 200 µl per well of 0.4% crystal violet solution to the adhered cells in the microtiter plates and left at room temperature for 15min. Excess stain was removed by rinsing the microtiter plates gently with 200µl of distilled water per well three times. These microtiter plates were then air-dried again for 1 hour and then 200µl of absolute ethanol was added to solubilize the dye. The intensity was measured at optical density (OD630nm) using a Safire Tecan-F129013 Micro Plate Reader (Tecan, Crailsheim, Germany). Background staining of each experiment was corrected by subtracting the crystal violet bound to un-inoculated controls from those of the samples using the formula by Omwenga *et al* (27). Antibiofilm formation inhibitory activity was carried out using the selected eight bacteria isolates (*E*.*coli* 3323, E.coli 5810, *Klebsiella spp* 3306, *Klebsiella spp* 3063, *Staphylococcus spp* 3309, *Staphylococcus spp* 3328, *Proteus spp* 3346, and *Proteus spp* 3268) that were obtained from our previous study (18). The treatment drugs used for the antibiofilm formation inhibitory activity assay were in different dosages/concentrations as follows; Ampicillin (AMP)-500mg/ml, 250mg/ml, 125mg/ml and 62.5mg/ml: Ciprofloxacin (CIP)-500mg/ml, 250mg/ml, 125mg/ml and 62.5mg/ml: Sulfamethoxazole (SXT) -960mg/ml, 480mg/ml, 240mg/ml and 120mg/ml: Ceftriaxone (CRO)-1000mg/ml,500mg/ml,250mg/ml and 125mg/ml. To estimate the antibiofilm activity (Abf A) of a given antibiotic, the following equation was used (27). *Antibiofilm formation ability (Abf A) (%) = (1-(ODTest sample - ODBlank)/ (OD Untreated sample – OD Blank) ×100*.

All these tests were carried out in triplicates independent of each other and *Pseudomonas aeruginosa* (ATCC-27853) was used as the study reference strain.

### 2.5. Data Analysis

The study findings were later entered into excels spread sheets for analysis using graph pad prism version. ANOVA Dunnett’s multiple comparisons test was applied to determine the antibiofilm formation inhibitory assays. Binary logistic regression analysis was carried out to generate the adjusted odds ratio with a 95% confidence interval. An alpha of less than 0.05 (P<0.05) was considered statistically significant.

## 3.0 Results

### 3.1 Study population and sample size

From the outpatient department, 174 (84.5%) were recruited, while inpatient 32 (15.5 %) were from the inpatients [18]. The overall age mean among study participants was 31.8 years. Out of the two hundred and six (206) midstream urine samples collected from the participants, only 57 had significant urinary tract infection growth, which translates to a prevalence of 27.6%. Therefore, the 57 significant isolates were used as the sample size of this current study and were subjected to EβSL and biofilm formation analysis (18).

#### 3.2.1 Prevalence of β-lactamase Phenotypes

From the 57 isolates that met the threshold for significant UTI, 14 (24.6%) were resistant to Ceftriaxone, Cefotaxime or Ceftazidime and were regarded as possible ESBL producers. However, only 10 (17.5%) were confirmed as ESBL producers by the double disk method (4).

Further screening for genotypic carriage of selected *bla* genes showed that TEM was present in all the 10 (100%) screened isolates revealing readings more than 867 bp, as demonstrated in **figure 3.2-C and Table 3.1** below. In addition, **Figure 3.2-A** demonstrates that EβSLs *bla*_*OXA*_ genes were detected 30% findings more than 854 bp among *E. coli* isolates while in **Figure 3.2-B** the prevalence of EβSLs genotype *bla*_*SHV*_ is demonstrated at 40% with findings more than 280 bp among *E. coli* isolates.

**Figure 3.1:**
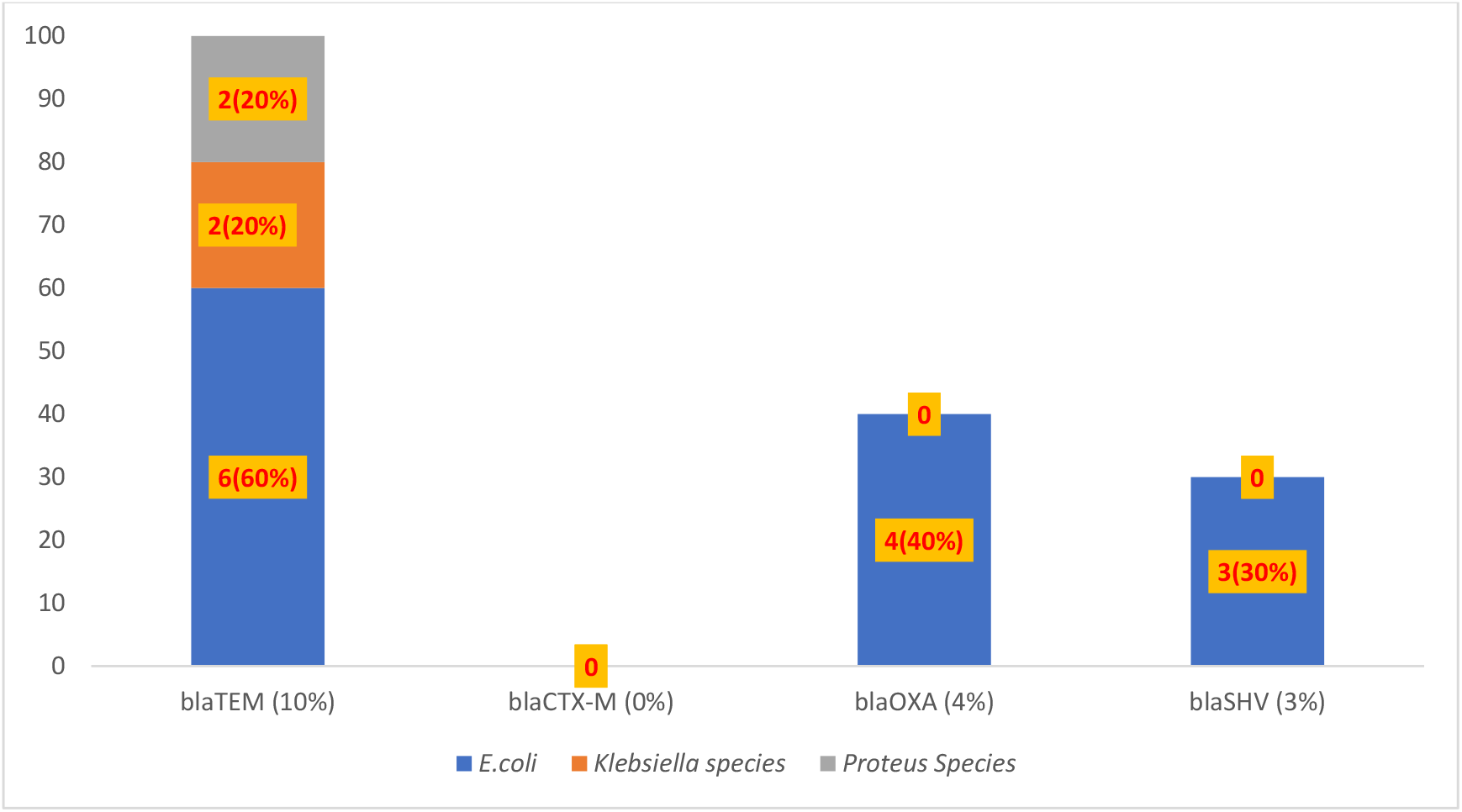
EβSLs genes results of the obtained bacterial isolates. **Key word: *bla***-*ß lactamase genes;* % - percentage

**Figure 3.2:**
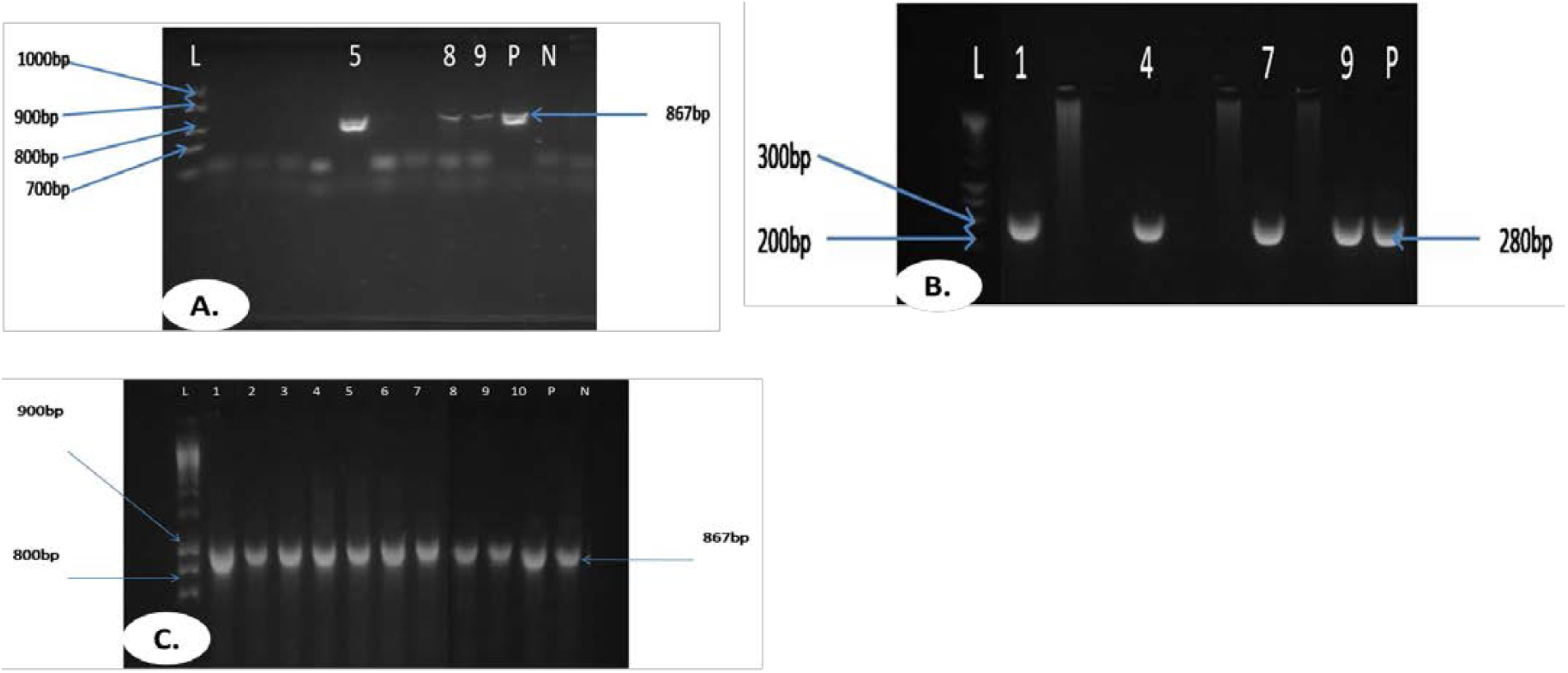
Electrophoresis gel results for *bla*_*SHV*_ genes (A), *bla*_*OXA*_ genes (B) and *bla*_*TEM*_ genes (C).

**Table 3.1.** EβSL genes co-carriage results and biofilm formation results.

| EβSL genes co-carriage (n) |  |  |  |  |
| --- | --- | --- | --- | --- |
| Organism | N | *bla*<sub>TEM</sub>+*bla*<sub>SHV</sub>+*bla*<sub>OXA</sub> | *bla*<sub>TEM</sub>+*bla*<sub>OXA</sub> | *bla*<sub>TEM</sub>+*bla*<sub>SHV</sub> |
| *E. coli* | 6 | 3(6) | 4(6) | 3(6) |
| *Klebsiella species* | 2 | 0(2) | 0(2) | 0(2) |
| *Proteus species* | 2 | 0(2) | 0(2) | 0(2) |
| All | 10 | 3 | 4 | 3 |

| Organism | Biofilm former | Non-biofilm formers | OR | 95% CI | P-value |
| --- | --- | --- | --- | --- | --- |
| *E. coli* | 15 | 9 | 1.03 | 0.384-2.758 | 0.9554 |
| *Klebsiella species* | 7 | 3 | 0.74 | 0.171-3.15 | 0.6781 |
| *Proteus species* | 3 | 3 | 1.71 | 0.317-9.276 | 0.5315 |
| *S. aureus* | 7 | 4 | 0.98 | 0.256-3.746 | 0.976 |
| *Enterococcus species* | 0 | 1 | 5.09 | 0.199-130.659 | 0.3254 |
| *S. saprophyticus* | 4 | 1 | 0.43 | 0.045-4.093 | 0.4617 |
| Gram- negative | 25 | 15 | 1.03 | 0.446-2.374 | 0.9474 |
| Gram-positive | 11 | 6 | 2.49 | 0.976-6.368 | **0.0561\*** |

| Department | Biofilm former-36(63.2%) | Non-biofilm former-21(36.8%) | Total (%) |
| --- | --- | --- | --- |
| Out patient | 32 | 17 | 49(85.9%) |

| In patient | 4 | 4 | 8(14 %) |
| --- | --- | --- | --- |
| Odds ratio |  |  | 1.860542 |
| P-value |  |  | 0.4053425 |
| 95% CI | 0.3057786 | 11.36315 |  |
| Gender | Biofilm former (%) | Non-biofilm former (%) | Total (%) |
| Men | 2(5.6%) | 1(4.7%) | 3(5.3%) |
| Female | 34(94.4%) | 20(95.2%) | 54(94.7) |
| odds ratio |  |  | 1.173229 |
| 95% CI | 0.05763235 | 72.80199 | 0.8970091 |

**Keyword:** *bla-* $\beta$ lactamase genes; % - percentage; N–number of cases

Analysis based on the co-carriage of EβSLs genes revealed the co-carriage of *bla*_*TEM*_ and *bla*_*OXA*_ among *E. coli* isolates at 66.7% and the co-carriage of *bla*_*TEM*_ + *bla*_*SHV*_ at (50%) as demonstrated on **Table 3.1** below.

### 3.3 Genetic diversity of E. coli isolates causing UTIs

The six isolates: Ec1-*E*.*coli* 5806; Ec2-*E*.*coli* 3352; Ec3-*E*.*coli* 3323; Ec4-*E*.*coli* 3177; Ec5-*E*.*coli* 3318 and Ec6-*E*.*coli* 3350 did not cluster based on similarities in antimicrobial resistance profiles or β-lactamases genes carriage. For instance, Isolates *E*.*coli* 5806 and *E*.*coli* 3350 had identical resistance profiles (AMP, CRO, CAZ, CTX) yet clustered in different clades. This was also similar to isolates *E*.*coli* 3352 and *E*.*coli* 3323 that also had identical antimicrobial resistance (AMP, ATM, CRO, CAZ, CTX) and clustered into two different clades. Therefore, the six isolates could be grouped into four clades with similarities of between 65 – 86%, which could reflect no indication of similar strain expansion in the Kiambu region, **Figure 3.3**.

**Figure 3.3:**
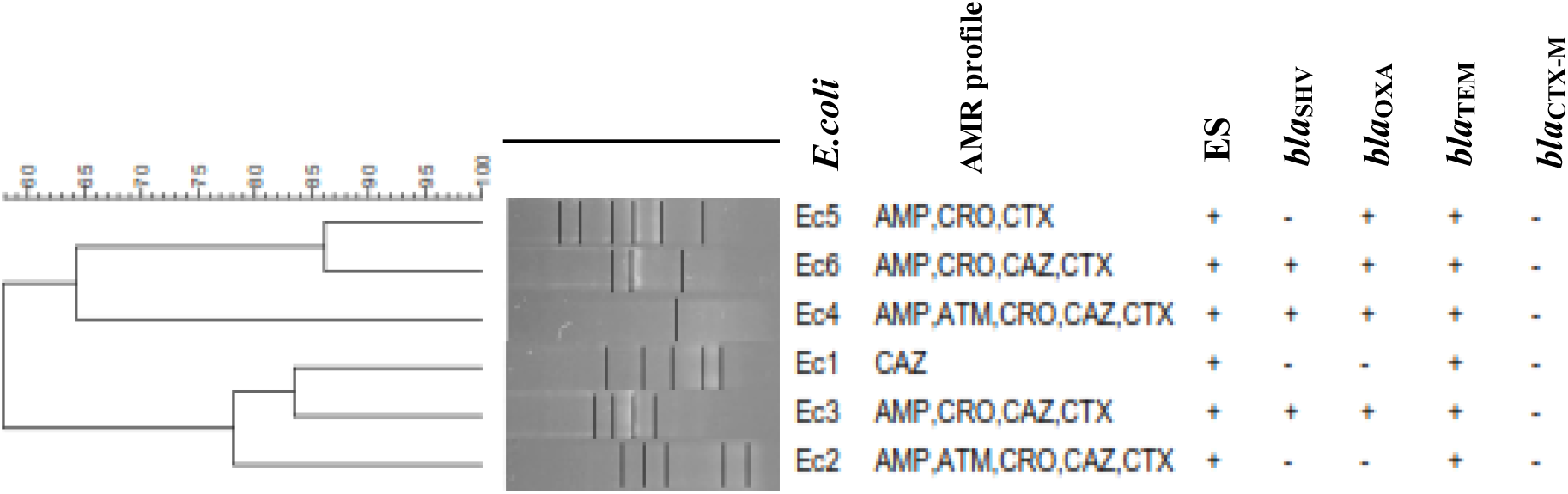
*Escherichia coli* dendrogram **Key:** The figure shows phylogeny analysis of *Escherichia coli* isolates that were positive for the carriage of EβSLs genes. Description of various acronyms on this figure is as follows; ***E. coli***: *Escherichia coli*, AMR: Antimicrobial resistance, *bla*_*TEM*_: Temoneria β-lactamase, *bla*_*CTX-M*_: Cefotaxime Munich β-lactamase, *bla*_*SHV*_: Sulhydryl variant β-lactamase, *bla*_OX_: *bla*_OXA_-type β-lactamases Figure 3.3.3 Dendrogram showing the genetic diversity of *E. coli* isolates.

#### 3.4.1 UTIs isolate Biofilm formation assay

The biofilm formation assays revealed that majority of the screened study isolates were biofilm formers - 63.2% (36/57) to non-biofilm formers at 36.8% (21/57). Gram-negative isolates had the highest biofilm formation prevalence at 69.5% (25/36) compared to the Gram-positive 30.5% (11/36). With *E. coli* 15(41.6%), *Klebsiella sp*. 7 (19.5%) and *S. aureus* 7 (19.5%) respectively being the most common biofilm formers as demonstrated in **Table 3.1 above**.

Further analysis based on significance of the results, showed no significant difference in biofilm formation among the tested isolates with a p-value of more than 0.05 recorded across board. However, analysis based on bacteria classification revealed that Gram positive isolates yielded significant biofilm formation ability with a notable P-value of 0.0561 **Table 3.1 above**.

From the 57 analysed isolates the outpatient department had 49 (85.9%), a prevalence of 88.9% (32/36) biofilm formers compared to the inpatient 11.1% (4/36). Non-biofilm formers prevalence findings were also similar with the outpatient biofilm formation prevalence at 80.9 % (17/21) to 19.1 % (4/21) of the in-patient department. In addition, biofilm formation was prevalent in women’s urine isolates at 62.9 % (34/57) compared to men’s at 5.6 % (2/57) as summarized in **Table 3.1 above**

#### 3.4.2 Antibiofilm formation inhibitory activity assay

Antibiofilm formation activity assay was carried out for eight bacteria isolates that were selected based on their antimicrobial resistance activity against test antibiotics in our previous study (18). The revealed data demonstrated that the biofilm formation inhibitory effects varied differently between the study isolates species and the various antibiotic dosages in comparison to the positive control as demonstrated in **Figure 3.5.1; 3.5.2, 3.5.3 and 3.5.4**. Ceftriaxone had the best antibiofilm inhibitory effects at all dosages compared to the rest of the test treatment antibiotics. On the other hand, Sulfamethoxazole yielded the least observed antibiofilm inhibitory effects across the four dosages as demonstrated in figure 3.5.3 below. Further analysis also revealed that Gram-negative isolates had the most antibiofilm inhibitory effects compared to the Gram-positive *Staphylococcus species*. In addition, analysis based on how each species were affected show *Proteus spp* were more susceptible to treatment drugs at various dosages compared to the rest of the isolates **Figure 3.5.1 to Figure 3.5.4**.

**FIG: 3.5.1.**
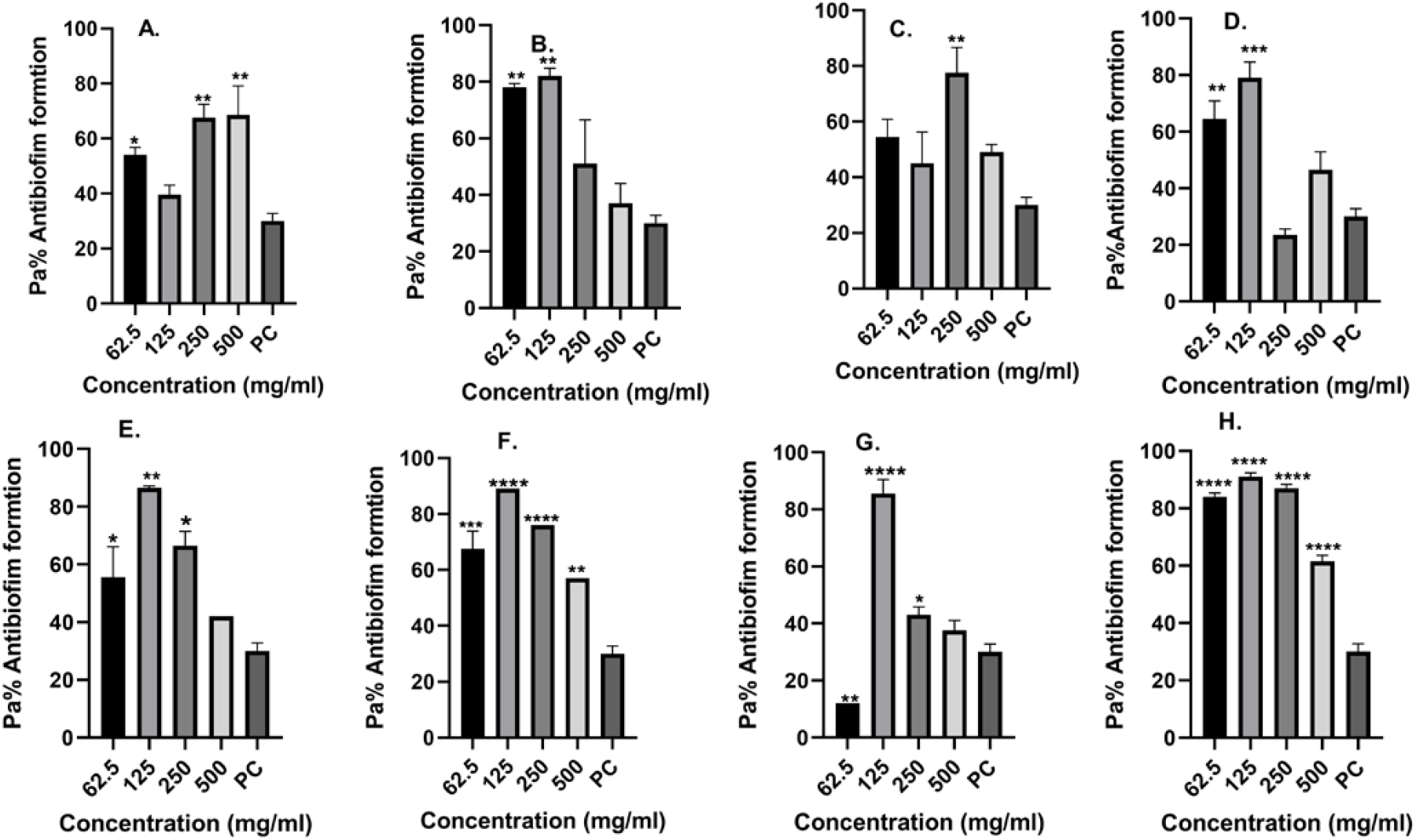
Ampicillin antibiofilm formation activity against (A) *E. coli* 3323 Isolate (B) *E. coli* 5810 Isolate (C) *Klebsiella spp* 3306 Isolate (D) *Klebsiella spp* 3063 Isolate (E) *Staphylococcus spp* 3309 Isolate (F) *Staphylococcus spp* 3328 Isolate (G) *Proteus spp* 3346 Isolate and (H) *Proteus spp* 3268 Isolate; PC=*P. aeruginosa* – Positive control (*n*=3, ANOVA Dunnett’s multiple comparisons test; \**P*=0.05; \*\**P*=0.01; \*\*\**P*=0.001; \*\*\*\**P*=0.0001).

**FIG: 3.5.2.**
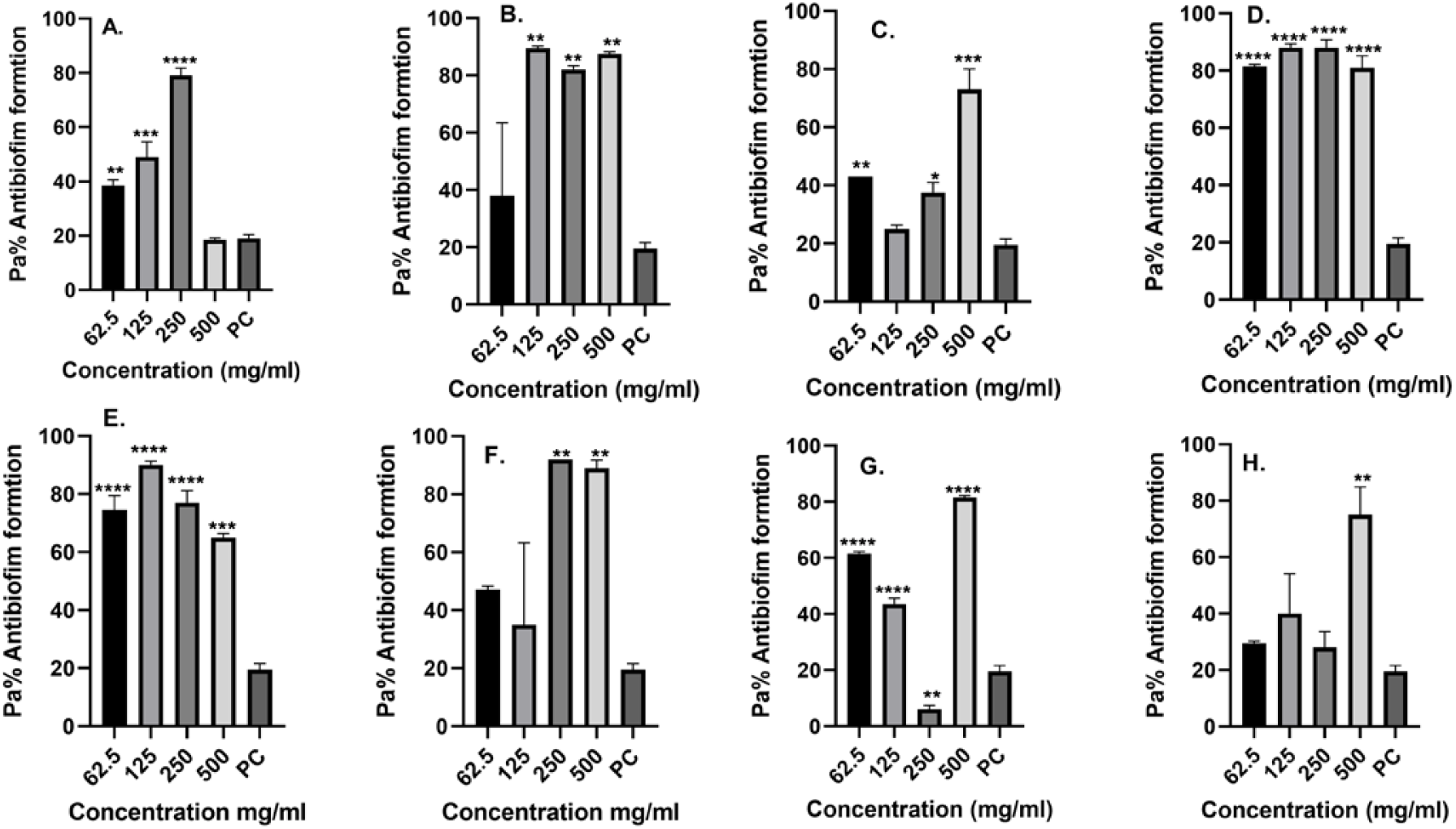
Ciprofloxacin antibiofilm formation activity against (A) *E. coli* 3323 Isolate (B) *E. coli* 5810 Isolate (C) *Klebsiella spp* 3306 Isolate (D) *Klebsiella spp* 3063 Isolate (E) *Staphylococcus spp* 3309 Isolate (F) *Staphylococcus spp* 3328 Isolate (G) *Proteus spp* 3346 Isolate and (H) *Proteus spp* 3268 Isolate; PC=*P. aeruginosa* – Positive control (*n*=3, ANOVA Dunnett’s multiple comparisons test; \**P*=0.05; \*\**P*=0.01; \*\*\**P*=0.001; \*\*\*\**P*=0.0001).

**FIG: 3.5.3.**
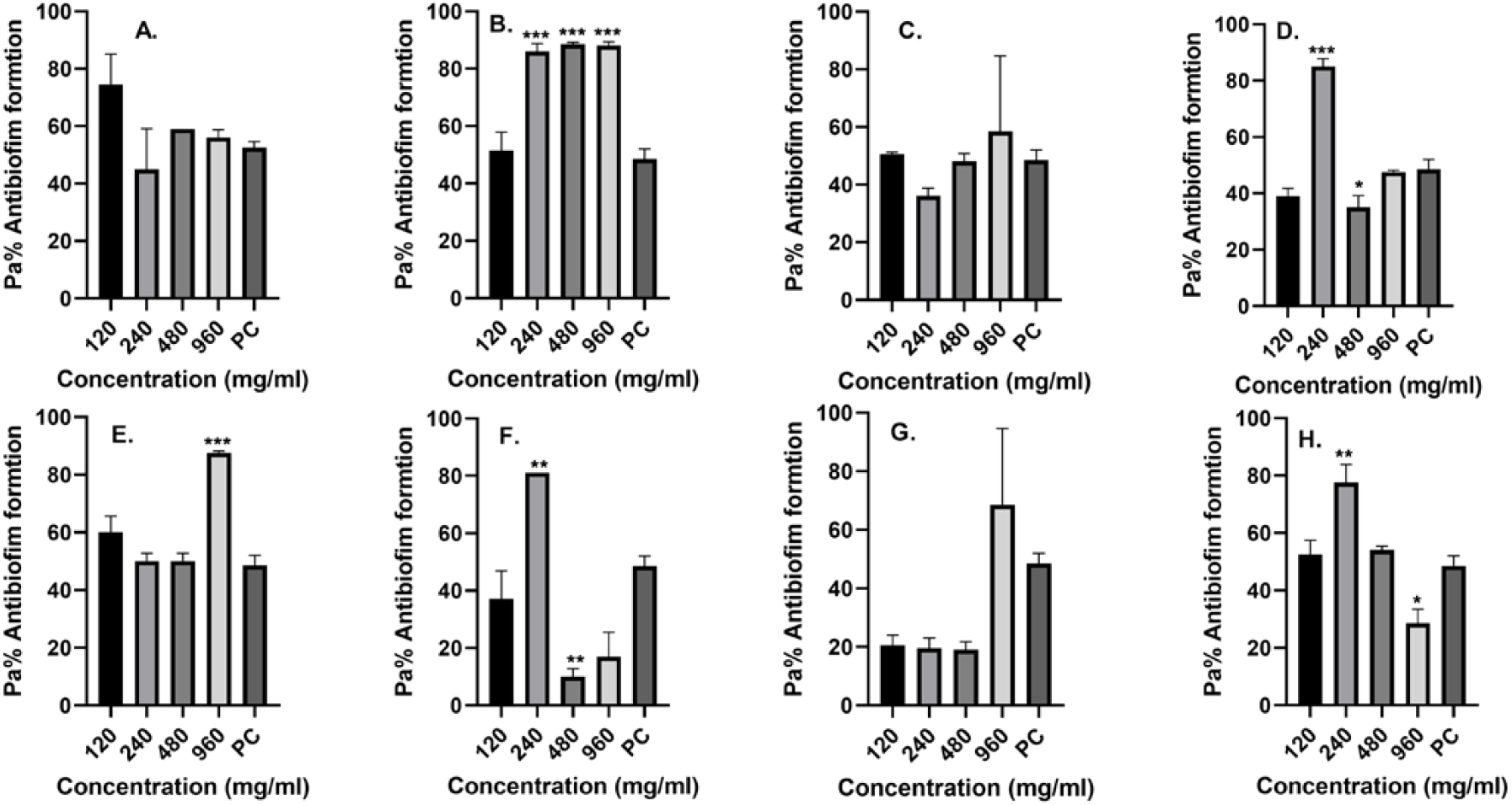
Sulfamethoxazole antibiofilm formation activity against (A) *E. coli* 3323 Isolate (B) *E. coli* 5810 Isolate (C) *Klebsiella spp* 3306 Isolate (D) *Klebsiella spp* 3063 Isolate (E) *Staphylococcus spp* 3309 Isolate (F) *Staphylococcus spp* 3328 Isolate (G) *Proteus spp* 3346 Isolate and (H) *Proteus spp* 3268 Isolate; PC=*P. aeruginosa* – Positive control (*n*=3, ANOVA Dunnett’s multiple comparisons test; \**P*=0.05; \*\**P*=0.01; \*\*\**P*=0.001; \*\*\*\**P*=0.0001).

**FIG: 3.5.4.**
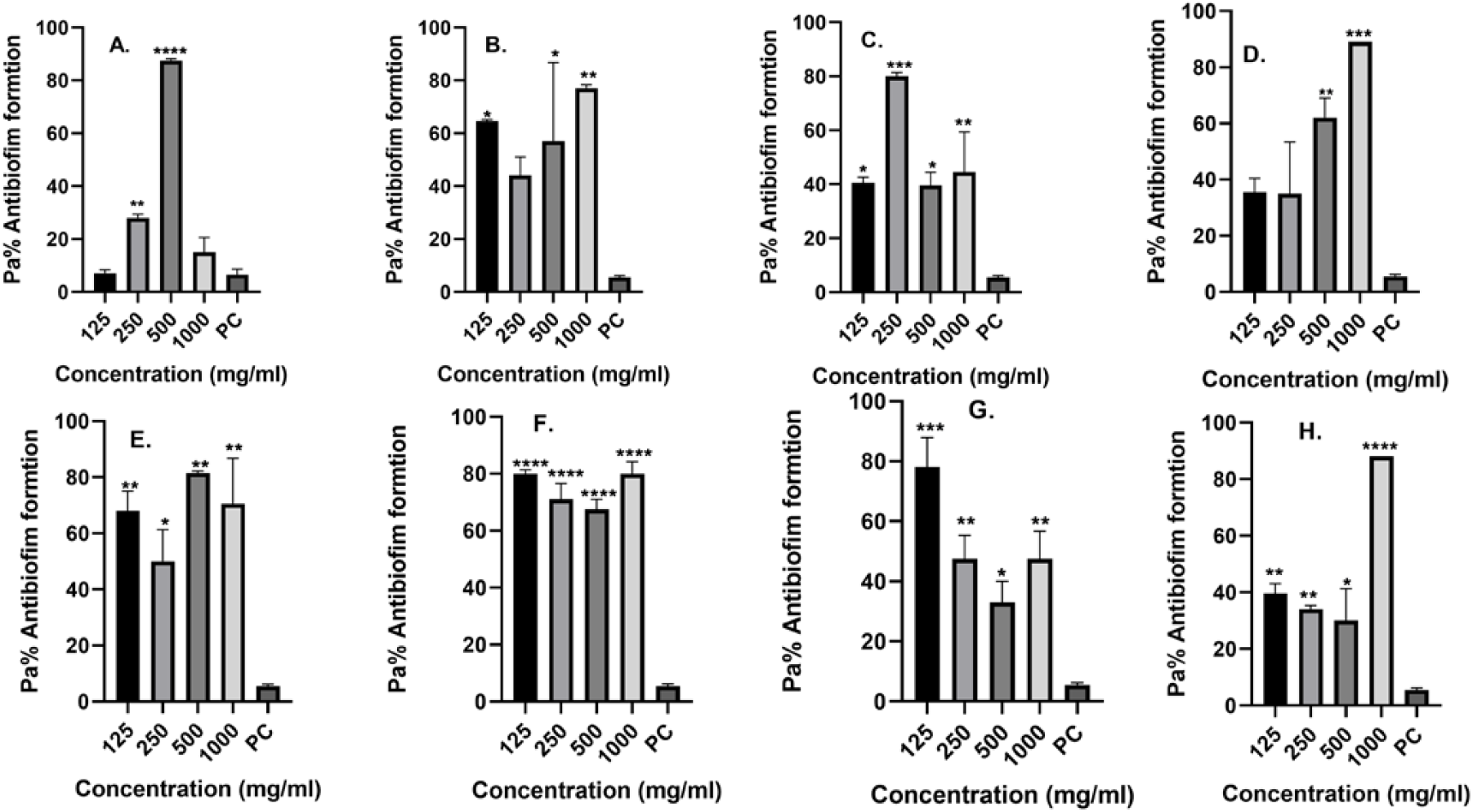
Ceftriaxone antibiofilm formation activity against (A) *E. coli* 3323 Isolate (B) *E. coli* 5810 Isolate (C) *Klebsiella spp* 3306 Isolate (D) *Klebsiella spp* 3063 Isolate (E) *Staphylococcus spp* 3309 Isolate (F) *Staphylococcus spp* 3328 Isolate (G) *Proteus spp* 3346 Isolate and (H) *Proteus spp* 3268 Isolate; PC=*P. aeruginosa* – Positive control (*n*=3, ANOVA Dunnett’s multiple comparisons test; \**P*=0.05; \*\**P*=0.01; \*\*\**P*=0.001; \*\*\*\**P*=0.0001).

##### 3.4.2.1: Ampicillin antibiofilm effect/activity

The antibiofilm effects findings against ampicillin revealed huge variation more so amongst isolates of the same species. For instance, among the *E. coli* isolates; *E. coli* 3323 isolate yielded moderate antibiofilm inhibitory effects especially when subjected to ampicillin 500mg/ml and 250mg/ml dosages when compared to that of *E. coli* 5810 isolate against the same 500mg/ml and 250mg/ml dosages **Figure 3.5.1 below**.

In addition, Staphylococcus *spp* 3328 and *Proteus spp* 3268 isolates in the various test drug dosages yielded significant antibiofilm formation inhibition activity (*P*= *** = 0.001; **** = 0.0001). However, across all the ampicillin dosages, *Klebsiella spp* 3306 yielded the least antibiofilm inhibition effect recording p value of ** = 0.01 against the dosage of 250 mg/ml. The revealed ampicillin antibiofilm inhibition findings are therefore promising and demonstrate that even though most of the isolates were resistant to ampicillin, it’s still a good antibiofilm formation drug.

Further analysis based on the ampicillin dosages inhibitory effects also demonstrated huge variations. The low dosages of 62.5 mg/ml and 125 mg/ml had significant differences on the antibiofilm inhibition activity compared to the higher dosages 250 mg/ml and 500mg/ml Figure 3.5.1 below. This observation may be explained by the fact at high concentrations sometimes aggregation of the drug can occur at the point of entry unto the bacteria cell and hence not much of the drug enters the target site.

##### 3.4.2.2: Ciprofloxacin antibiofilm effect/activity

The antibiofilm inhibitory finding against ciprofloxacin showed majority of the isolates being significantly affected when subjected to the various ciprofloxacin dosages. For instance, *Klebsiella spp* 3063 and *Staphylococcus spp* isolates were greatly affected revealing significant antibiofilm inhibitory effects in presence of the different ciprofloxacin test dosages (p value of **** = 0.0001). However, moderately antibiofilm inhibitory activities were yielded among isolates *E. coli* 3323; *E. coli* 5810 and *Klebsiella spp* 3306 recording p value of ** = 0.01 and *** = 0.001respectively.

The least affected isolates were the *Proteus spp* isolates with *Proteus spp* 3268 isolate yielding no inhibitory effect when subjected to 62.5mg/ml, 125 mg/ml and 250 mg/ml ciprofloxacin dosages as compared to its counterpart *Proteus spp* 3346 isolate that was highly affected **Figure 3.5.2 below**.

Further antibiofilm inhibitory findings based on the actual inhibitory impact among the different ciprofloxacin dosages also revealed the 250mg/dl dosage having the highest inhibitory effects. This was closely followed by 62.5mg/ml and 125 mg/ml dosages respectively, while the 500mg/ml dosage had the least inhibitory effects.

##### 3.4.2.3: Sulfamethoxazole antibiofilm effect/activity

The inhibitory analysis of sulfamethoxazole dosages against the study isolates yielded the least of inhibitory effect overall compared to the rest of the study test drugs as demonstrated in **Figure 3.5.3 below**. However, sulfamethoxazole inhibitory effect analysis based on how the isolate species were affected revealed the *E. coli* isolates as the most affected species. *E. coli* 5810 isolate particularly was the most affected and recorded the highest antibiofilm formation activity especially when subjected to sulfamethoxazole dosages of 240 mg/ml, 480 mg/ml and 960 mg/ml; p value of *** = 0.001. In addition, the study *Klebsiella* 3063 isolate yielded moderate inhibitory effects compared to *Klebsiella spp* 3306 isolate that had no inhibitory effects recorded at all. Moderate inhibitory activity was also revealed by *Proteus spp* 3268 isolates with a p value of * = 0.05 compared to the other Proteus isolate *Proteus spp* 3346 that had no inhibitory effects.

More analysis based on the various test drugs dosages inhibitory performance also revealed the lower dosages performed better than the higher dosages. However, over all the 240mg/ml dosage performed better than the rest of the Sulfamethoxazole dosages as demonstrated in **Figure 3.5.3 below**.

##### 3.4.2.4: Ceftriaxone antibiofilm effect/activity

Ceftriaxone was the most effective test drug according to our study antibiofilm inhibitory activity analysis. All the ceftriaxone dosages had high or moderate inhibitory effects against all the eight study isolates. However, analysis based on how individual isolate was affected revealed *Staphylococcus species* were the most affected compared to the rest of the study isolates species. *Staphylococcus spp* 3328 isolate was the most highly affected yielding an antibiofilm inhibitory activity of a p value of **** = 0.0001compared to the other *Staphylococcus spp* 3309 isolate that had been moderately affected. Further analysis based on how also demonstrated that most of the other isolates had moderately antibiofilm formation activity with the only significant variation seen being that of *Klebsiella spp* 3063 that recorded no significant inhibitory effect when subjected to 125mg/ml and 250 mg/ml dosages.

In concurrence to the rest of the study test drugs, ceftriaxone dosages also had minimum inhibitory effect yielded by the very high test dosages of 500mg/ml and 1000mg/ml dosages at p values of ** = 0.01 and p value of \**P*=0.05 respectively compared to the lower dosages of 125mg/ml and 250 mg/ml **Figure 3.5.4 below**.

## 4.0 Discussion

Bacterial biofilms and carriage of resistance genes may be the basis of most established and persistent urinary tract infections cases. Though scientific research has indicated that there is an ever-increasing antimicrobial resistance and biofilm formation in most infectious diseases causing pathogens including the urinary tract infection causative agents, more studies are needed to tackle this public health issue. Findings from the present study demonstrated the landscape of the interplay between biofilm formation and antimicrobial resistance among the study’s isolates. Considering the high UTI prevalence rate of 27.6% and the phenotypic resistance profiles to commonly used antibiotics reported in our previous study (18). It is plausible that the carriage of EβSLs genes may be the major driving factor that influenced resistance of the study isolates against the test antibiotics. The carriage of *bla*_*TEM*_ was the most prevalent, detected at 100% (10/10) closely followed by *bla*_*OXA*_ genes at 40% (4/10) and *bla*_*SHV*_ at 30% (3/10) respectively. This signals a clear indication that *bla*_*TEM*_ genes may have greatly impacted the isolates resistance profiles as this gene is responsible for resistance towards important classes of ß-lactams antibiotics like Cefuroxime, Cefotaxime and Ceftriaxone as reported in previous studies (28)(29).

Findings from our previous study on the presence of *bla*_*TEM*_ genes seemed to imply that the genes enhanced the ability of obtained bacteria isolates to inactivate ampicillin at a range of 17% to 67%, Cefotaxime at 18% to 50 and Ceftriaxone at 18% to 29% (18). Moreover, the carriage of *bla*_*TEM*_ genes among *Klebsiella* and *Proteus* species was recorded at 20% respectively; an observation that may be afflicted to the revealed high resistance profiles of *Klebsiella* and *Proteus* species against Sulfamethoxazole and Ampicillin at range between 50% - 85% (18).The revealed *bla*_*TEM*_ finding therefore concurs with other findings from related studies that documented *bla*_*TEM*_ genes being responsible for increasing bacteria pathogen’s hydrolysing activity against penicillin and first-generation Cephalosporin (28)(29).

On the other hand, the carriage of *bla*_*SHV*_ and *bla*_*OXA*_ genotypes among *E. coli* and *Klebsiella pneumoniae* isolates was also revealed at 40% and 30 % respectively, a confirmation that the distribution of these EβSL genes have spread across most of the commonly known bacteria of medical importance. These two EβSL genotypes have in the recent past been documented to being able to confer drug and multidrug resistance (29), with the most commonly reported significant resistance being noticed against extended-spectrum cephalosporins, a class of antibiotics that are of immense clinical significance (29). This observation therefore underpins the role of the carriage of *bla*_*SHV*_ and *bla*_*OXA*_ EβSL genotypes as major contributors of the rising treatment failure. However, going by this study data, more surveillance program to monitor the carriage of new mutant *bla*_*SHV*_ and *bla*_*OXA*_ genes that are emerging is urgent (30). This is so because *bla*_*SHV*_ genes are often encoded by self-transmissible plasmids and can easily be transmitted along with other resistance genes leading to significant clinical challenges (4). Furthermore, the current epidemiolgy of *bla*_*SHV*_ genes have also shown that their prevalence among members of the Enterobacteriaceae have been on the rise in the last decade, and that the genes have evolved from a narrow to an extended-spectrum of hydrolzing activity,especially against monobactams and carbapenems antibiotic classes (31).

In the current study, the carriage of *bla*_*OXA*_ gene was actually common among the study *E. coli* isolates, and going by past reported findings this *bla*_*OXA*_ genotype is associated with wide range of difficult-to-treat infections owing to their hydrolyzing activity towards a variety of antibiotics (32). This therefore may have influenced the revealed findings where most of the *E. coli* obtained isolates expressed drug and multidrug resistance trends towards test antibiotics that are commonly used in the Kenyan health facilities to treat UTI. This included resistance towards ceftazidime, cefepime, nitrofurantoin and much more against ampicillin at a range between 60-75% (33). However, the occurrence of strains that did express co-carriage of resistance genes still raises lots of health concerns more so if they are implicated in infections, since there will be very minimal available treatment options.

The carriage of *bla*_*CTX-M*_ EβSL genotype among uropathogens have been recorded to have led to major health challenges in the treatment of community acquired UTIs (4), this study however differs a lot with the others studies since the presence of carriage *bla*_*CTX-M*_ EβSL genotype was not detected. The *bla*_*CTX-M*_ genes have been documented as the most commonly known ESβLs genotype obtained among uropathogens that is easily acquired via horizontal gene transfer (29)(34). In addition, these *bla*_*CTX-M*_ genes are known to be of significant medical challenge as they enable bacterial pathogens to easily hydrolyze Cefotaxime one of the many commonly used antibiotic to treat UTIs (29). Even so, the luck of *bla*_*CTX-M*_ isolation in the study demonstrates there might be a new dynamic of the distribution of resistance genes among uropathogens and based on that our data, we advocate for routine screening of the carriage of EβSL genes to help inform and guide the periodic revision of UTI management. Furthermore, the obtained genotypic characterization finding of uropathogens will also be needed to obtain data that will aid in reducing the rate of spread and emergence of new resistance genes. Though the total cessation of emergence and spread of AMR genes still seems to be impossible, it’s upon the health stakeholders to accelerate education campaigns on UTIs as a prophylactic measure to enlighten the society and assist halt UTI contraction.

This study phylogeny analysis showed significant similarity among the *E. coli* isolates at more than 70%. The *E. coli* strains had the same identical resistance phenotypes and shared more than 75% genetic similarity. Though previously documented finding reported a positive correlation between genetic diversity and acquisition of antimicrobial resistance among bacteria uropathogens (35), this study finding is different and varies with that of a previous study where *E. coli* isolates exhibited high rates of genetic change as well as exhibition of high resistance rates against test antibiotics (34). However, the finding is of significant clinical importance since it reveals how some strains of the same genetic similarity can possess drug and multi-drug resistance genes. Close monitoring, and screening of genetic similarity among uropathogens is therefore paramount to aid in management and halt spread of pathogens that have become more notorious to treat by a range of clinically known antimicrobial agents (35).

In addition, the formation of biofilms by pathogenic bacteria and the contribution of these biofilms to AMR is also another situation that is complex and causes a significant health dilemma. For example, the gains and losses of resistance genes and the ability to form biofilms by bacteria especially those causing urinary tract infection need to be known to shed light on more crucial knowledge of the dynamics that occur within the said biofilms. According to the past documented findings, positive correlation between biofilm-related genes and antibiotic resistance genes is a largely notable impact that affects the static/cidal actions of antibiotics (10),(12),(36). Microbes inside these said biofilms are protected from attack by the host immune system or even the administered antibiotics leading to disease establishment and enhanced treatment failure (10), (12), (36). From this study majority of the isolates had the ability to form biofilm-at 63.2% and although our data analysis showed no significant difference in biofilm formation among all tested isolates with the p-values being more than 0.05, it is worth noting that overall, the Gram-positive isolates had significant P-value of 0.0561 **Table 3.1**. Gram-negative isolates were the most prevalent in forming biofilms at 69.5% (25/36) compared to the Gram-positive 30.5% (11/36). An observation that may be afflicted to the high resistance profiles finding shown by Gram-negative isolates against test drugs; Ampicillin, Cefotaxime, Cefepime and Ciprofloxacin at range of more than 40 % (18). Further analysis based on our comparative biofilm formation between Gram-negatives and Gram-positives isolates, revealed *E. coli* isolates still as the most prevalent biofilm formers at 41.7 % (14). The biofilm reduces the microbes’ susceptibility to antibiotics due to the biofilm exopolysaccharide polymers formed by the mass of bacterial cells that houses them. This allows the microbes more time to keep on multiplying and producing more virulence traits. The biofilm ability observation seen among this study *E. coli* isolates coupled with the carriage of *blaTEM, blaSHV* and *bla*_*OXA*_ EβSL genotypes underpins the exemplary high antimicrobial and multidrug resistance trends revealed among the *Escherichia coli* isolates towards Sulfamethoxazole, Nalidixic acid and Ampicillin of 64%, 50% and 27% respectively (18). This may be a revelation as to why *E. coli* in particular is becoming more lethal and more difficult pathogen to treat. The finding is in agreement with a related study that termed *E. coli, S. aureus* and *P. aeruginosa* as the most notorious biofilm formers isolated from hospital and community-based infections (8). The other Gram-negative biofilm formers from the current study included *Klebsiella* and *Proteus* isolates at 19.4% (6) and 8.3% (9) respectively.

For the Gram-positive study isolates, *S. aureus* isolates was the most prevalent biofilm formers at 19.4% (6), followed by S. *saprophyticus* at 11.1% (10). The overall findings on biofilm formation ability by both Gram-negatives and Gram-positives isolates therefore revealed a logical explanation that most uropathogens are able form biofilms, that in return enable them to easily proliferate in the urinary tract since the biofilm protect them from the acidic environment and the potent effects of the host immune responses and other antimicrobials. Additionally, these biofilms allow easy transfer of resistance genes by the cells that are closely placed and eventually may be playing an important role of facilitating plasmid horizontal gene exchange between closely positioned microbes and enhancing the spread of resistance genes (37).

Our biofilm formation analysis based on gender also revealed isolates obtained from women samples being more of biofilm formers at 62.9% (35) compared to men at 5.6% (4). This finding is of significant importance and could be attributed to several factors like women’s anatomy and indwelling intrauterine contraceptive device that partly enhances the establishment of biofilms by being instrumentation harbour or act as reservoirs of uropathogens. These provide anchorage support for microbial biofilm formation and also hinder easy flushing out of the attached uropathogens by urine (35); however, more research should be done to ascertain this assumption.

In the current study, the ability of test drugs to inhibit biofilm formation activity was also analyzed. This was done using selected eight bacterial isolates: *E. coli* 3323, *E. coli* 5810, *Klebsiella spp* 3306, *Klebsiella spp* 3063, *Staphylococcus spp* 3309, *Staphylococcus spp* 3328, *Proteus spp* 3346, and *Proteus spp* 3268 based on their antimicrobial resistance trends in our previous study (18). The antibiofilm formation activity revealed significant findings of antibiofilm inhibitory effects across the different antibiotic’s dosages; Ampicillin, Ceftriaxone, Ciprofloxacin and Sulfamethoxazole **Figure; 3.5.1; 3.5.2; 3.5.3 and 3.5.4**. These results are in tandem with those from a past study (27) and are still in agreement to the antimicrobial susceptibility profiles in our previously conducted study (18). We can therefore anticipate that use of this drugs as a prophylactic measure against uropathogens may be a future solution to counter uropathogens microbial biofilm formation and eventually halt spread of resistance genes that have largely led to the rising AMR. However, this suggestion needs a feasibility study to be fully validated.

Further antibiofilm inhibitory activity analysis revealed that test treatment drugs; Ampicillin, Ciprofloxacin and Ceftriaxone dosages yielded the most significant antibiofilm inhibitory effects against the screened isolates recording p values above 0.05 (*). On the other hand, Sulfamethoxazole had the least antibiofilm inhibitory effects, an observation that concurs with the observed antimicrobial susceptibility test result where most the isolates exhibited high resistance trends against Sulfamethoxazole at a range between 40% and 80% (18). Ceftriaxone had the highest antibiofilm inhibitory effects which may be afflicted to the drugs mode of action since it inhibits the mucopeptide synthesis of bacteria cell wall thus preventing bacteria cell wall and cell division (10). However, Sulfamethoxazole mode of action is different as it involves inhibition of dihydropteroate synthesis enzyme to prevent for bacterial synthesis of purines and DNA. This mode of action cannot prevent the initial bacteria growth hence affords the bacteria ample time to form biofilms (28). Even though understanding how antibiotics may inhibit biofilm formation is crucial, it should also be seen starting point to understanding the interplay between biofilm formation and the AMR. Health stake holders should therefore fund more health campaign rallies and give out handouts on biofilm education to enlighten the public domain on the ease of spread and acquisition of antimicrobial resistance genes within biofilms. This will increase the limited knowledge in the society and enlighten people on how easily resistance genes are transferred specifically those attributed to plasmid-borne integrons that is able to assemble genes in cassettes resulting to compromised disease treatment (15)(38).

Further analysis based on the antibiofilm inhibitory effects demonstrated that in most of the test antibiotics, less inhibitory effects were seen at high dosages as compared to lower ones. This is not an abnormal finding as it has been documented that at high dosages sometimes aggregation can occur of the drug at the point of entry unto the bacteria cell and hence not much of the drug enters the target site. This for instance, was documented by Omwenga *et al*., (27), when they were working with nanoencapsulated flavonoids against quorum sensing and Awour *et al*., (39) when they were working with antibiofilm formation of *V. cholerae* isolates against selected antibiotics. Nevertheless, several solutions have been identified to counteract biofilms formation such as rifampicin for Gram-positive bacteria and fluoroquinolones for Gram-negative bacteria (40), the available anti-biofilms options are however still limited and leave a huge health concern gap. More options are needed to increase the available choices and intensify the fight against antimicrobial resistance trends since most of the antibiofilm compounds are not bacteria cidal and only render the cells in a planktonic growth state, which are more susceptible to antibiotics and more easily cleared by the immune system (20). Scientists should lobby and embrace more biofilm research work to open up more channels for exploring new anti-biofilm formation inhibitory strategies and curb the contribution of biofilms in microbial survival, persistence and encouragement antibiotic resistance.

## 5.0 Conclusion

According to the current study, the ultimate goal to achieve amicable antimicrobial stewardship and curb the rising treatment failure of urinary tract infection could be via a holistic campaign of mapping the carriage of resistance genes and biofilm formation. This will help revise the existing empiric treatment regimens now informed by the data of the prevailing resistance phenotypes. Besides, the data will assist in preserving the potency of β-lactam antibiotics and improve success of chemotherapy. The public domain will again be reassured the of a health solution, since there is limited hope of new novel drugs.

However, this study was only able to screen a few selected ESBLs genes and the possibility of biofilm formation among the screened bacteria etiological agents. Future studies that will focus more on detection of other AMR genes against other class antibiotics and biofilm genes are encouraged. The development of novel antibiotics, biofilm formation inhibitory compounds and vaccines against bacteria causing urinary tract infections will also be a milestone achievement for the modern medicine.

## 6.0 Study Limitations

This study had a number of shortcomings that can be addressed in future surveillance studies;

1. We were only able to for a few EβSLs genotypes. Future studies to determine the carriage of other β-lactamase genes are fundamental. We were also not able to screen the carriage of other classes of resistance genes that are associated to resistance to other classes of antimicrobial agents like fluoroquinolones and aminoglycosides.
2. We were only able to determine the possibility of biofilm formation among the screened isolates. However, future studies can therefore involve the screening of carriage of genes associated with biofilms to shed lighter on bacterial host invasion and pathogenesis.

## Data Availability

All data is made available in the manuscript.

## Acknowledgements

To the study participants, thank you for agreeing to take part in this study. The authors wish to further express heartfelt gratitude to all KEMRI/CMR (molecular biology) staff members particularly Susan Wambui Kiiru for their invaluable support. We are also grateful to the clinicians, laboratory technologists, and the administration Kiambu level 5 Hospital for their guidance and assistance. This study was funded by HATUA (Holistic Approach to Unravel Antimicrobial resistance in East Africa) research funds. The funders had no role in study design, data collection and analysis, decision to publish, or manuscript preparation.

## Author’s Contribution

Fredrick Wanja (FW) was the principal investigator and came up with the study concept. FW designed, carried out the lab work, data analysis and drafted the manuscript. John Maina participated in sample collection, lab work and drafting of the manuscript. Dr. Caroline Ngugi and Dr. Eric Omwenga assisted in preparation and harmonization of the manuscript. Dr. John Kiiru took part in the study design and supervised the lab bench work. All authors read and endorsed the manuscript.

## Author’s Information

Fredrick Kimunya (FK) is a master’s student in Kenya studying Infectious Diseases and Vaccinology at Jomo Kenyatta University of agriculture and technology. John Maina (JM) is a scientific researcher at the Centre of Microbiology Research, Kenya Medical Research Institute, Nairobi, Kenya. Dr. Caroline Ngugi (CN) is the Chairperson Department of Medical Microbiology, Jomo Kenyatta University of agriculture and technology who possess vast experience in proposal development. Dr. Eric Omwenga (EO) is the Chairperson Department of Microbiology & Parasitological, Kisii University. He assisted in coming up with the study concept and revision of the content of the manuscript. Dr. John Kiiru (JK) a senior researcher at the Centre of Microbiology Research, Kenya Medical Research Institute, Nairobi, Kenya and the Head of Laboratory Ministry of Health, Nairobi, Kenya who have vast knowledge in infectious diseases and microbiology took part in the study design and actualization of the content of the manuscript.

## Ethical Approval

Approved (Supplementary data **S** 1 and Supplementary data **S** 2)

## Supplementary Data S1

### Purpose of the research study?

Dear participant you are requested to take part in this study. The study aims to obtain information about the genotypic characterization and biofilm formation of isolated uropathogens among adult patients attending Kiambu level 5 hospital. Kindly read through the following information and also listen as I explain to you all that the study entails. In case of any words, terms, or sections that are not clearly understood, you are highly encouraged to seek further explanation. Ask any question about the study that you feel it concerns you. If you accept to take part in the study, please fill in this form and keep your copy for personal records. The form has important information about the names and physical address which you can use in the future to make any inquiries about the study.

### You are invited to consent or decline to consent to do the following

1. Provide a urine sample
2. Allow us to keep the urine specimen obtained from you by freezing or any other means of long term preservation.
3. Allow storage and use of bacterial isolate strains from this study for future studies related to UTI investigations or further analysis.

### I would also like to disclose to you the following information involving the study

- Some of the questions you are about to be asked may be uncomfortable or personally, however, you are assured that optimum privacy and confidentiality before consenting, during and post the study exercises will be observed.
- Data collected will only be accessed by the study relevant authorities and only used for the purposes of this study.
- You may find it a bit challenging or embarrassing to collect urine sample but I will explain to you the best and most comfortable manner to do so.
- The main purpose of obtaining a urine sample from you is to determine whether you have developed a UTI or not. Isolated uropathogens will further be subjected to antimicrobial susceptibility test to determine appropriate drug of choice. Genetic relatedness of the isolates will also be determined to make good informed study conclusions.
- There are no incentives or monetary gains for participating in this study.
- Participation in this study is on a voluntary basis and one can decline to continue to answer or participate if you feel like.
- One can also withdraw from the study any time you feel like and this will not draw any penalties or discrimination in the future.
- My role as the principal investigator is to provide all the information about the study and the findings data to the clinicians at Kiambu level 5 hospital to aid in treating you. However, you will be required to cater for bills that will arise from your sickness like purchasing the prescribed antibiotics.

### Your role as the participant in this research study

You will be required to provide a urine sample that will be used for investigation of UTI causing bacteria. For the purpose of the study, you are also required to consent on your behalf, after which you will now obtain a urine sample. The urine sample will not be stored after culture is performed but disposed according to KEMRI biosafety procedures immediately. Please note that the isolated biological samples may be used for future studies and you are therefore requested to consent to allow their use as and when required.

### Additional procedure for patients who will participate in this study

For urine samples taken and found to be positive of UTI, the participant results will be shared with the clinicians who will then treat you. Sample collection will be on sterile universal container and you will be given instructions on how to best collect and pack the clean catch mid-stream urine sample. You will collect the urine sample within the hospital compound and provide it to the lab immediately. Only samples provided to the lab within two hours will be accepted.

### What will happen after the study?

Data obtained from this study will be used to determine the various bacterial species prevalence, antimicrobial susceptibility profiles, biofilm formation ability and carriage of ESBLs genes by the isolated bacterial uropathogens among adult patients attending Kiambu level 5 hospital. This data will also provide critical information that will enhance the diagnosis of these infections in the facility and aid in treatment of participants found to be positive of UTI. The data will help to enlighten the facility administration if there is need to revise and improve policies on how to manage UTI among patients visiting the facility. This is in regard to conducting routine urine culture and prescription of antibiotics.

### What happens in case the participant has a change of mind?

If you had agreed to take part in the study and with time you happen to change your mind, you are free to withdraw at any time with no penalties. You will also not be discriminated in any way in the future due to your decision to withdraw.

### Who will read or hear about information collected from the participant

The information collected from the participant will be stored using the assigned study number/ code since no name will be indicated. The coded information will also be stored in soft copy format on computers protected by passwords only known by the study relevant authorities.

### Do you have any questions about the study that you feel was left out, was not addressed or was not well understood?

Feel free to ask and I will answer you now. If you would like to inquire more details about the research study or have any issues about your rights to participate that need to be discussed in the future, you can contact any of the following people;

1. **The Secretary, KNH-UoN Ethics and Research Committee. College of Health Sciences P.O Box 19676-00202, Nairobi Tel. (254-0200) 2726300-9 Ext 44355**.
2. **Fredrick Kimunya Wanja (Principal Investigator) -+254724089245**.
3. **Dr. John N Kiiru, CMR-KEMRI - +254721805285**
4. **Dr. Caroline Ngugi, Dept. of Medical Microbiology, JKUAT - +254722556790**
5. **Dr. Eric Omwenga, Dept. of Bio &Nanotechnology, Kisii University- +254725806875**

**Laboratory number…………………………………………**.

**Telephone number……………………………………………**

I have read the above information to the best of my knowledge and have had the opportunity to ask questions which were all answered to my satisfactory. I consent to participate in the study as has been explained and as I have understood it.

**Signature …………………………………**.

**Date …………………………………**.

Right or Left hand thumb print for those who cannot sign

**Name of principle investigator……………………………………………………………**

**Signature……………………………… Date…………………………………**.

